# The Progressive Supranuclear Palsy Clinical Deficits Scale (PSP-CDS) tracks longitudinal progression in PSP

**DOI:** 10.64898/2026.07.23.26358755

**Authors:** Jun Ping Chua, Tzi Shin Toh, Darren Wan Pin Lim, Ting Qing Lim, Kelly Kher Nie Chen, Yi Xuan Tee, Yi Zhe Lim, Jeremy Christopher Fernandiz, Kah Hui Yap, Yi Wen Tay, Hans Xing Ding, Anis Nadhirah Khairul Anuar, University of Malaya PSP Study Group, Global Parkinson’s Genetics Program (GP2), Ai Huey Tan, Hirotaka Iwaki, Shen-Yang Lim

**Author notes:** Co-First authors (Jun Ping Chua, Tzi Shin Toh). University of Malaya PSP Study Group.

## Abstract

**Background:** The Progressive Supranuclear Palsy Clinical Deficits Scale (PSP-CDS) is a brief rating scale of clinical severity in PSP. However, its longitudinal performance has not been evaluated. We aimed to assess the ability of the PSP-CDS to track disease progression over time and to compare progression across PSP phenotypes in a real-world Asian cohort.

**Methods:** Patients who met the Movement Disorder Society PSP diagnostic criteria and underwent at least 2 PSP-CDS assessments were recruited from movement disorders clinics in Malaysia. Longitudinal progression was evaluated using linear mixed-effects models. Domain-specific progression and subtype-specific trajectories were also analyzed. Associations between annualized changes in PSP-CDS and Barthel Index (BI) scores were examined.

**Results:** 104 patients (including 59 with PSP-Richardson’s syndrome [PSP-RS], 28 with predominant parkinsonism [PSP-P], and 14 with progressive gait freezing [PSP-PGF]) contributed 394 PSP-CDS assessments over a median follow-up of 33.2 months (range, 8.7– 73.1 months). PSP-CDS scores increased significantly over time (β=0.126 points/month), corresponding to estimated increases of 1.13 points over 9 months and 2.27 points over 18 months. Subtype-specific analyses demonstrated the fastest progression in PSP-RS (0.156 points/month), followed by PSP-PGF (0.081 points/month) and PSP-P (0.077 points/month). Exploratory domain-level analyses showed that finger dexterity, communication, and dysphagia were the most rapidly worsening domains. Annualized PSP-CDS progression correlated significantly with annualized decline in BI scores (Spearman’s ρ=-0.474, P<0.001).

**Conclusions:** The PSP-CDS is sensitive to longitudinal disease progression in PSP and captures clinically-meaningful functional decline. Its brevity and ability to distinguish differential progression across PSP phenotypes support its utility as a pragmatic outcome measure for routine clinical practice and research.

## INTRODUCTION

Accurate assessment of clinical disease severity is fundamental in movement disorders practice and research, to assess disease severity cross-sectionally, and to monitor progression and treatment response over time. Yet, in the realm of Parkinsonian disorders, patient assessments often lack detailed phenotypic characterization and, particularly, quantification of disease severity [1]. In large part, this is because the available gold-standard instruments to perform such assessments (e.g., of motoric and cognitive function) are often time-consuming and laborious for busy clinicians, as well as for patients and caregivers who may have other pressing priorities, or are limited by advanced disease (e.g., dementia), fatigue, apathy, or mood and anxiety disorders [2,3].

Thus, there is currently an unmet need for practical and brief, yet accurate, clinical rating scales [2,3]. In an effort to address this gap in progressive supranuclear palsy (PSP), the International Parkinson and Movement Disorder Society (MDS) PSP Study Group created the PSP Clinical Deficits Scale (PSP-CDS), which captures seven functionally relevant clinical aspects of the disease [4]. Building upon this, we demonstrated the feasibility of the PSP-CDS in a real-world setting, which, taking only ∼5-10 mins to complete, could be applied to consecutive PSP patients seen in routine clinical practice [5,6]. The scale also showed strong correlations with functional disability, as measured by the Barthel Index (BI), and with patient- and caregiver-reported outcomes [6].

To date, however, no study has been published on the characteristics of the PSP-CDS when used to track PSP longitudinally. Moreover, prospectively-collected data on the longitudinal progression of PSP patients in underrepresented populations, including Asians, are very scarce [7]. Currently, understanding of the differential progression patterns in the classic Richardson’s syndrome (PSP-RS) vs. the “subcortical” forms of PSP (with predominant parkinsonism [PSP-P] or progressive gait freezing [PSP-PGF]) is derived entirely from European-ancestry populations [8–10]. We therefore aimed to study the longitudinal performance of the PSP-CDS in a cohort of Malaysian patients with PSP, encompassing PSP-RS and variant subtypes.

## METHODS

### Study Population and Clinical Assessments

Patients with PSP were recruited from movement disorders clinics at the University of Malaya Medical Centre (UMMC) and University of Malaya Specialist Centre (UMSC). PSP diagnosis and phenotypic classifications were done by experienced movement disorders neurologists (SYL and AHT) according to the MDS PSP clinical diagnostic criteria [11], as previously described [5]. Clinico-demographic data, including age, sex, ethnicity, age at symptom onset, age at first PSP-CDS assessment, and disease duration, were collected. Longitudinal disease severity was assessed using the PSP-CDS (in its original English version), administered by SYL and AHT during routine follow-up visits and repeated whenever ≥9 months had elapsed since the previous PSP-CDS rating. Formal permission to use the scale was granted by the MDS on 5 October 2022.

Functional status was evaluated using the BI and repeated after ≥18 months, as time permitted in the clinic (i.e., convenience sampling). The study was approved by the institutional ethics review board (MREC ID numbers: 20191010-7917 and 202515-14559), and all patients or their proxies provided written informed consent.

### Statistical Analysis

Statistical analyses were performed using R version 4.6.0. Continuous variables are presented as median [interquartile range (IQR)] or mean ± standard deviation (SD), as appropriate. Categorical variables are presented as frequencies and percentages. Comparisons across PSP phenotypes were performed using one-way analysis of variance (ANOVA) for normally distributed variables, the Kruskal–Wallis test for non-normally distributed variables, and the Chi-square test for categorical variables.

Longitudinal progression of PSP-CDS scores was evaluated using linear mixed-effects models fitted with the lmer function from the lme4 package [12]. Follow-up duration was defined as the interval between the first and last available PSP-CDS assessments for each participant. The primary model included time from baseline (months), baseline PSP-CDS score, age at first PSP-CDS assessment, sex, and PSP phenotype as fixed effects. To account for individual variability in disease progression, participant-specific random slopes for follow-up time were included: PSP-CDS ∼ Time_months + Baseline_CDS + Age_baseline + Sex + PSP_subtype + (0 + Time_months | ID). Models were fitted using restricted maximum likelihood. Type III analysis of variance with Satterthwaite’s approximation for degrees of freedom was used to evaluate the significance of fixed effects. Subtype-specific analyses were subsequently performed for the three most common PSP phenotypes (PSP-RS, PSP-P, and PSP-PGF) using the same modelling framework, excluding PSP phenotype as a covariate. Exploratory analyses of individual PSP-CDS domains were performed in the overall cohort and within each PSP phenotype. Baseline-adjusted linear mixed-effects models incorporating participant-specific random slopes for follow-up time were fitted for each domain: Domain ∼ Time_months + Baseline_domain_score + Age_baseline + Sex + (0 + Time_months | ID), where the baseline domain score corresponded to the baseline value of the domain being analyzed where the baseline domain score referred to the baseline value of the corresponding domain (e.g., baseline communication score for the communication model). Estimated annual progression rates (β/year) were obtained by multiplying the monthly regression coefficients by 12.

Clinical trial sample size estimates were calculated assuming a 30% reduction in PSP-CDS progression, 90% statistical power, a two-sided significance level of α=0.05, and equal allocation between treatment groups. Estimates were derived using subtype-specific progression rates and corresponding residual standard deviations obtained from the fitted linear mixed-effects models. Additional analyses were performed to evaluate the impact of different participant retention rates on estimated sample size requirements.

Associations between annualized changes in PSP-CDS and BI scores were assessed using Spearman’s rank correlation coefficient. As BI assessments were not always performed contemporaneously with PSP-CDS assessments, the nearest available PSP-CDS assessment was matched to each BI assessment for correlation analyses. Temporal separation between BI and PSP-CDS assessments was evaluated to assess the robustness of this matching approach.

Statistical significance was defined as P<0.05 (two-tailed).

## RESULTS

### Cohort Characteristics

Of the 180 patients with PSP in the overall cohort, 104 had at least two PSP-CDS assessments and were included in the longitudinal analyses. These included 59 patients with PSP-RS, 28 with PSP-P, 14 with PSP-PGF, and 3 with other PSP variants (Table 1). A total of 394 PSP-CDS assessments were available for longitudinal analysis. The median age at the first PSP-CDS assessment was 72.1 years (IQR 65.9–76.5 years), and the median baseline PSP-CDS score was 9.0 (IQR 7.0–12.0). The median follow-up duration was 33.2 months (IQR 22.0–49.0 months; range 8.7–73.1 months). Participants underwent a median of 3.5 PSP-CDS assessments (IQR 3–5). Baseline characteristics of patients included in and excluded from the longitudinal analyses are summarized in Supplementary Table 1. The two groups were comparable with respect to sex, ethnicity, age at symptom onset, age at baseline, disease duration, baseline PSP-CDS score, and PSP subtype (all P>0.05).

**Table 1:** Clinico-demographic characteristics of the study cohort.

|  | Overall | PSP Subtypes |  |  |  | P-value |
| --- | --- | --- | --- | --- | --- | --- |
|  |  | PSP-RS | PSP-P | PSP-PGF | Other PSP Variants |  |
| **N** | 104 | 59 (56.7) | 28 (26.9) | 14 (13.5) | 3 (2.9) | - |
| **Sex** |  |  |  |  |  | 0.013* |
| Male | 61 (58.7) | 30 (50.8) | 23 (82.1) | 7 (50.0) | 1 (33.3) |  |
| Female | 43 (41.3) | 29 (49.2) | 5 (17.9) | 7 (50.0) | 2 (66.7) |  |
| **Ethnicity** |  |  |  |  |  | 0.892 |
| Chinese | 73 (70.2) | 41 (69.5) | 20 (71.4) | 10 (71.4) | 2 (66.7) |  |
| Indian | 20 (19.2) | 13 (22.0) | 4 (14.3) | 2 (14.3) | 1 (33.3) |  |
| Malay | 9 (8.7) | 4 (6.8) | 3 (10.7) | 2 (14.3) | 0 (0.0) |  |
| Others | 2 (1.9) | 1 (1.7) | 1 (3.6) | 0 (0.0) | 0 (0.0) |  |
| **Age at symptom onset** (years) | 67.0 [61.0-72.0] | 68.0 [62.5-73.0] | 66.0 [58.8-71.2] | 64.0 [63.0-68.5] | 59.0 [58.5-63.0] | 0.194 |
| **Age at baseline** (years) | 72.1 [65.9-76.5] | 72.3 [67.0-76.6] | 72.1 [65.1-77.3] | 69.5 [66.6-75.0] | 63.0 [60.9-67.9] | 0.508 |
| **Disease duration at baseline** (years) | 4.5 [2.6-6.4] | 3.4 [2.0-5.0] | 6.9 [5.1-8.8] | 5.6 [2.7-7.8] | 4.0 [2.4-4.9] | <0.001* |
| **PSP-CDS at baseline** | 9.0 [7.0-12.0] | 9.0 [7.0-12.0] | 10.0 [8.0-12.0] | 6.0 [5.0-7.0] | 8.0 [6.0-9.5] | 0.010* |
| **No. of follow-up assessments** | 2.5 [2.0-4.0] | 2.0 [2.0-3.0] | 2.5 [1.0-4.0] | 4.0 [2.2-4.0] | 3.0 [3.0-3.5] | 0.177 |
| **Follow-up duration** (months) | 33.2 [22.0-49.0] | 29.9 [21.7-41.1] | 33.1 [23.2-53.4] | 46.2 [36.0-61.4] | 41.5 [36.9-44.9] | 0.088 |
| **Barthel Index at baseline** | 65.0 [30.0-90.0] | 52.5 [26.2-85.0] | 47.5 [31.2-65.0] | 92.5 [77.5-95.0] | 70.0 [57.5-80.0] | 0.123 |

Categorical variables are presented as n (%), and continuous variables are presented as median [Q1–Q3].
P-values were calculated using Pearson's chi-square test for categorical variables and the Kruskal–Wallis rank-sum test for continuous variables.
\*Denotes statistically significant differences (P<0.05).
PSP=Progressive supranuclear palsy; PSP-CDS=PSP Clinical Deficits Scale; PSP-PGF=PSP with progressive gait freezing; PSP-P=PSP with predominant parkinsonism; PSP-RS=PSP Richardson's syndrome.

### Longitudinal Progression of PSP-CDS

As illustrated in Figure 1, substantial inter-individual variability in longitudinal PSP-CDS trajectories was observed; however, the overall trend demonstrated progressive worsening over time. In the primary baseline-adjusted linear mixed-effects model, PSP-CDS scores increased significantly over time (β=0.126 points/month, SE=0.013, 95% CI 0.101–0.151, t =9.72, P<0.001) (Table 2). Based on the estimated monthly progression rate, PSP-CDS scores increased by approximately 1.13 points over 9 months and 2.27 points over 18 months. Baseline PSP-CDS score was significantly associated with longitudinal PSP-CDS scores (P<0.001), whereas neither age at baseline nor sex was significantly associated with longitudinal PSP-CDS progression. PSP subtype was not significantly associated with longitudinal PSP-CDS scores in the overall model (Type III ANOVA, P=0.775).

**Figure 1:**
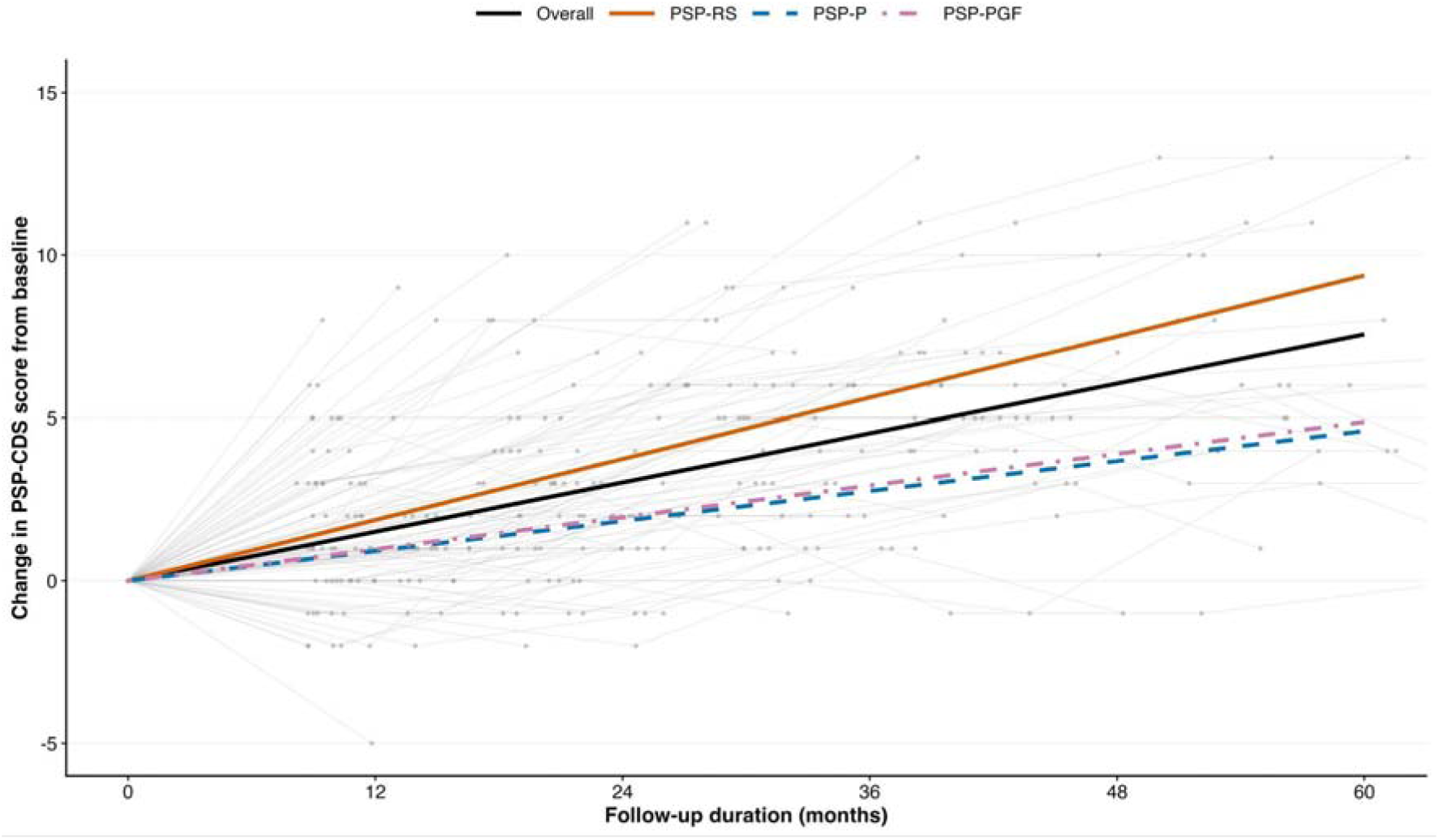
Longitudinal progression of PSP-CDS scores in the overall cohort and across major PSP subtypes. Grey lines and points represent individual participant trajectories. Coloured lines represent fitted trajectories estimated from baseline-adjusted linear mixed-effects models for the overall cohort (black; 0.126 points/month, 394 observations from 104 participants), PSP-RS (orange; 0.156 points/month, 211 observations from 59 participants), PSP-P (blue dashed; 0.077 points/month, 107 observations from 28 participants), and PSP-PGF (purple dot-dash; 0.081 points/month, 63 observations from 14 participants). Models were adjusted for baseline PSP-CDS score, age at baseline, sex, and (for the overall model) PSP subtype. PSP-RS demonstrated the fastest estimated rate of disease progression over time.

**Table 2:** Linear mixed-effects model of longitudinal PSP-CDS progression in the overall cohort and across PSP subtypes.

| Group | Measure | $\beta$ (SE) | 95% CI | T-value | P-value |
| --- | --- | --- | --- | --- | --- |
| Overall cohort<br>(n = 104) | Time from baseline (months) | 0.126 (0.013) | 0.101 to 0.151 | 9.72 | <0.001 |
|  | Baseline PSP-CDS score | 0.927 (0.029) | 0.870 to 0.984 | 31.90 | <0.001 |
|  | Age at baseline (years) | -0.003 (0.015) | -0.031 to 0.026 | -0.18 | 0.856 |
|  | Male sex (reference: Female) | -0.221 (0.216) | -0.645 to 0.203 | -1.02 | 0.308 |
|  | PSP subtype (reference: PSP-RS) | Overall Type III test | - | - | 0.775 |
| PSP-RS<br>(n = 59) | Time from baseline (months) | 0.156 (0.021) | 0.115 to 0.198 | 7.35 | <0.001 |
|  | Baseline PSP-CDS score | 0.956 (0.030) | 0.897 to 1.016 | 31.63 | <0.001 |
|  | Age at baseline (years) | 0.009 (0.017) | -0.024 to 0.042 | 0.54 | 0.590 |
|  | Male sex (reference: Female) | -0.278 (0.225) | -0.719 to 0.162 | -1.24 | 0.217 |
| PSP-P<br>(n = 28) | Time from baseline (months) | 0.077 (0.018) | 0.042 to 0.112 | 4.32 | <0.001 |
|  | Baseline PSP-CDS score | 0.880 (0.099) | 0.686 to 1.075 | 8.87 | <0.001 |
|  | Age at baseline (years) | -0.023 (0.031) | -0.084 to 0.038 | -0.74 | 0.461 |
|  | Male sex (reference: Female) | -0.099 (0.651) | -1.376 to 1.177 | -0.15 | 0.879 |
| PSP-PGF<br>(n = 14) | Time from baseline (months) | 0.081 (0.016) | 0.051 to 0.112 | 5.21 | <0.001 |
|  | Baseline PSP-CDS score | 0.909 (0.061) | 0.791 to 1.028 | 14.99 | <0.001 |
|  | Age at baseline (years) | 0.023 (0.040) | -0.054 to 0.101 | 0.59 | 0.558 |
|  | Male sex (reference: Female) | -0.552 (0.465) | -1.463 to 0.360 | -1.19 | 0.241 |

Linear mixed-effects models included follow-up time (months), baseline PSP-CDS score, age at baseline, sex, and PSP subtype (overall model only) as fixed effects. Participant-specific random slopes for follow-up time were included to account for between-participant variability in disease progression ((0 + Time\_months | ID)). P-values were estimated using Satterthwaite's approximation. Regression coefficients represent the estimated monthly change in PSP-CDS score after adjustment for model covariates.
$\beta$ = regression coefficient; CI = confidence interval; PSP-CDS = Progressive Supranuclear Palsy Clinical Deficits Scale; PSP-PGF=PSP with progressive gait freezing; PSP-P=PSP with predominant parkinsonism; PSP-RS=PSP Richardson's syndrome. SE = standard error.

### Progression According to PSP Phenotype

Subtype-specific analyses demonstrated significant progression across all three major PSP phenotypes (Table 2). PSP-RS exhibited the fastest progression rate, with an estimated monthly increase of 0.156 points (SE=0.021, P<0.001), corresponding to a model-derived increase of 1.41 points over 9 months. PSP-PGF and PSP-P demonstrated slower progression rates of 0.081 and 0.077 points/month, respectively (SE=0.016 and 0.018; both P<0.001), corresponding to model-derived 9-month increases of 0.73 and 0.69 points, respectively. As an exploratory analysis, the expected cumulative PSP-CDS progression was compared with the residual standard deviation estimated from the subtype-specific baseline-adjusted linear mixed-effects models. The residual standard deviation was used as an approximate threshold for detectable progression, representing the point at which the expected cumulative disease-related change exceeded typical unexplained model variability. Using this approach, the estimated time required for cumulative PSP-CDS progression to exceed the residual variability was shortest in PSP-RS (8.1 months), followed by PSP-PGF (15.9 months) and PSP-P (16.5 months) (Figure 2). As this threshold has not been formally validated as a measure of detectable clinical progression, these findings should be interpreted as exploratory.

**Figure 2.**
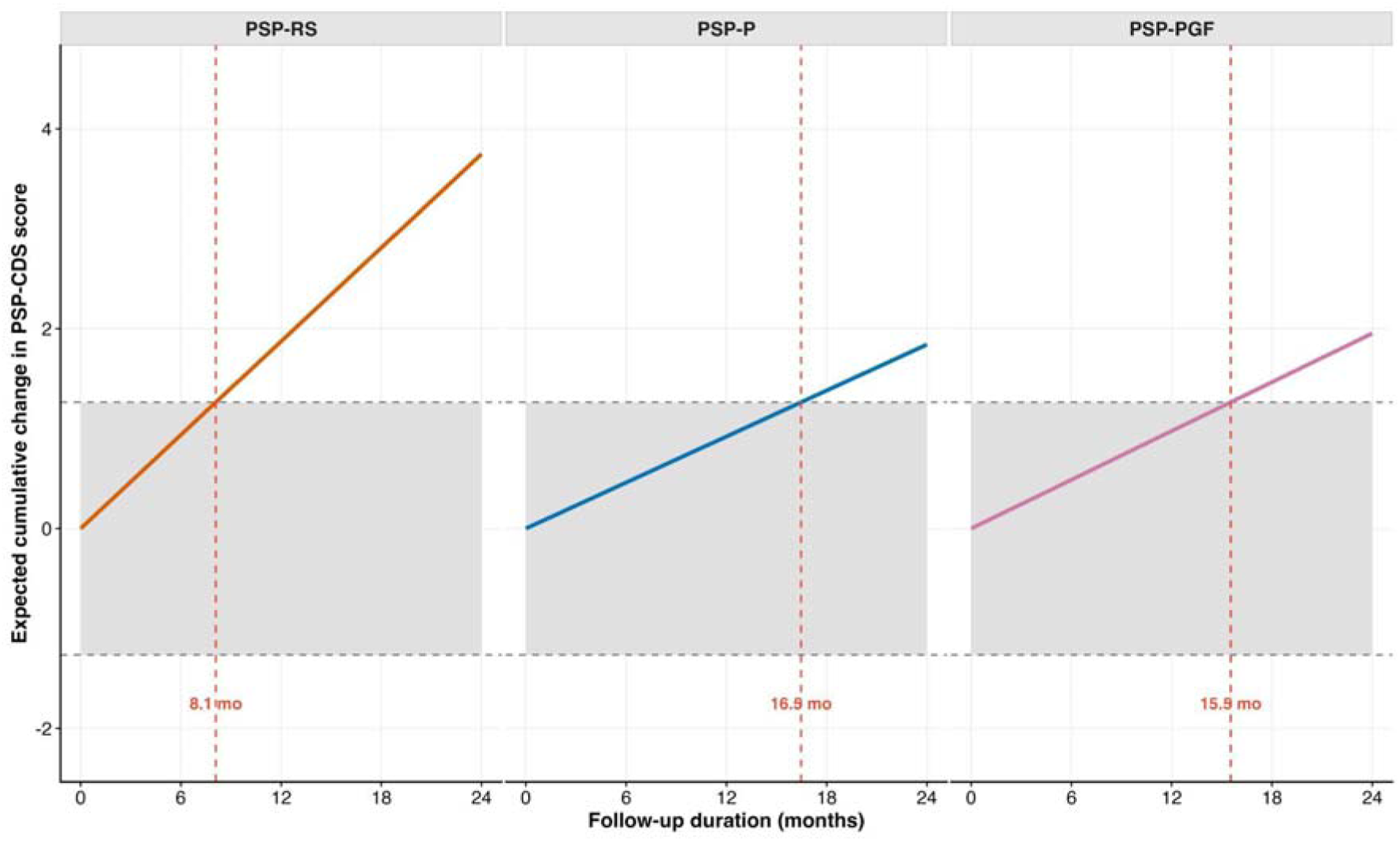
Estimated time required for cumulative PSP-CDS progression to exceed residual variability. Expected cumulative PSP-CDS progression was estimated from subtype-specific baseline-adjusted linear mixed-effects models. The shaded regions represent ±1 subtype-specific residual standard deviation, reflecting the unexplained within-participant variability remaining after accounting for baseline PSP-CDS score, age, sex, and participant-specific random slopes. The residual standard deviation was used as an approximate threshold for detectable progression, representing the point at which the expected cumulative disease-related change exceeds the typical magnitude of unexplained measurement variability. Vertical dashed lines indicate the estimated time required for the expected cumulative PSP-CDS change to exceed this threshold. Earlier threshold crossing indicates greater sensitivity for detecting longitudinal disease progression.

### Progression of Individual PSP-CDS Domains

Exploratory analyses of individual PSP-CDS domains demonstrated longitudinal worsening across all seven domains. Finger dexterity showed the greatest estimated annual progression (β = 0.264 points/year), followed by communication (β = 0.262 points/year), dysphagia (β = 0.244 points/year), gait and balance (β=0.226 points/year), and eye movements (β=0.221 points/year). Bradyphrenia (β = 0.145 points/year) and akinesia-rigidity (β = 0.093 points/year) demonstrated slower rates of progression. Supplementary Table 2 summarizes the estimated annual progression across individual PSP-CDS domains. Exploratory subtype-specific domain analyses demonstrated distinct progression patterns across PSP phenotypes, with PSP-RS generally exhibiting the greatest deterioration across finger dexterity, communication, dysphagia, and eye movement domains. Given the multiple statistical comparisons performed across domains and PSP subtypes, these findings should be interpreted as exploratory and hypothesis-generating. Detailed domain-level analyses are presented in Figure 3, Supplementary Figure 1 and Supplementary Table 2.

**Figure 3:**
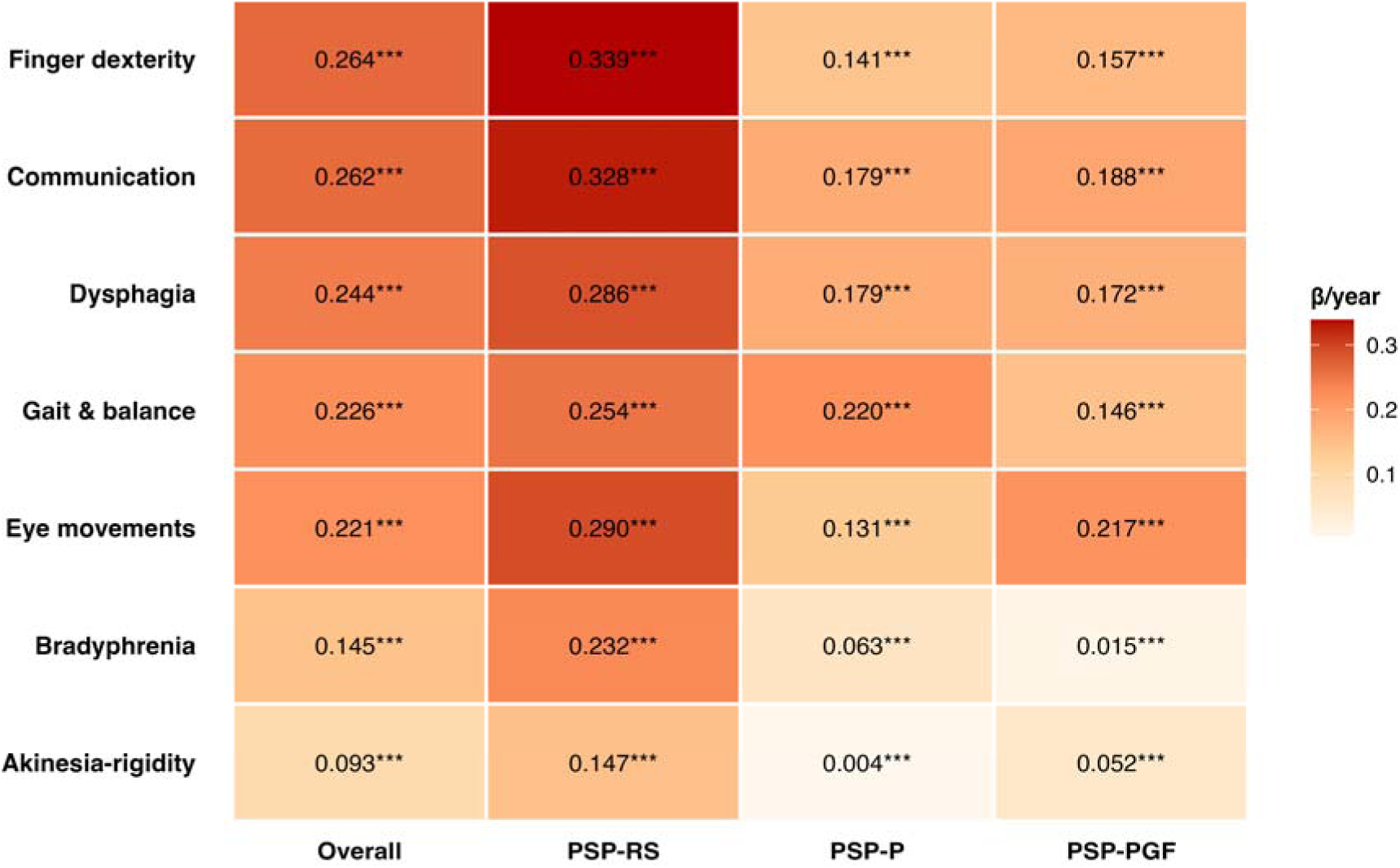
Estimated annual progression of individual PSP-CDS domains across major PSP subtypes. Heatmap illustrating estimated annual changes (β/year) in individual PSP-CDS domains derived from baseline-adjusted linear mixed-effects models for the overall cohort and subtype-specific analyses. Colour intensity reflects the magnitude of estimated annual progression, with darker colours indicating greater worsening. Numerical values represent estimated annual progression (β/year), with statistical significance for the effect of follow-up time indicated by asterisks (*P<0.05, **P <0.01, ***P<0.001). Domains are ordered according to the magnitude of annual progression observed in the overall cohort to facilitate comparison across the overall cohort and PSP subtypes..

### Association between PSP-CDS Progression and Functional Decline

To evaluate the functional relevance of PSP-CDS progression, annualized changes in PSP-CDS scores were correlated with annualized changes in BI scores (Figure 4). Longitudinal BI data suitable for correlation analyses were available for 54 patients. A moderate inverse correlation was observed between annualized changes in PSP-CDS and BI score (Spearman’s ρ=-0.474, 95% CI 0.693 to −0.216, P<0.001), indicating that patients with faster PSP-CDS progression experienced greater declines in functional independence. Temporal alignment between BI and PSP-CDS assessments was high, with 48 of 54 (88.9%) baseline assessment pairs and 41 of 54 (75.9%) follow-up assessment pairs performed on the same day (Supplementary Table 3), supporting the validity of the matching approach used for the correlation analyses.

**Figure 4:**
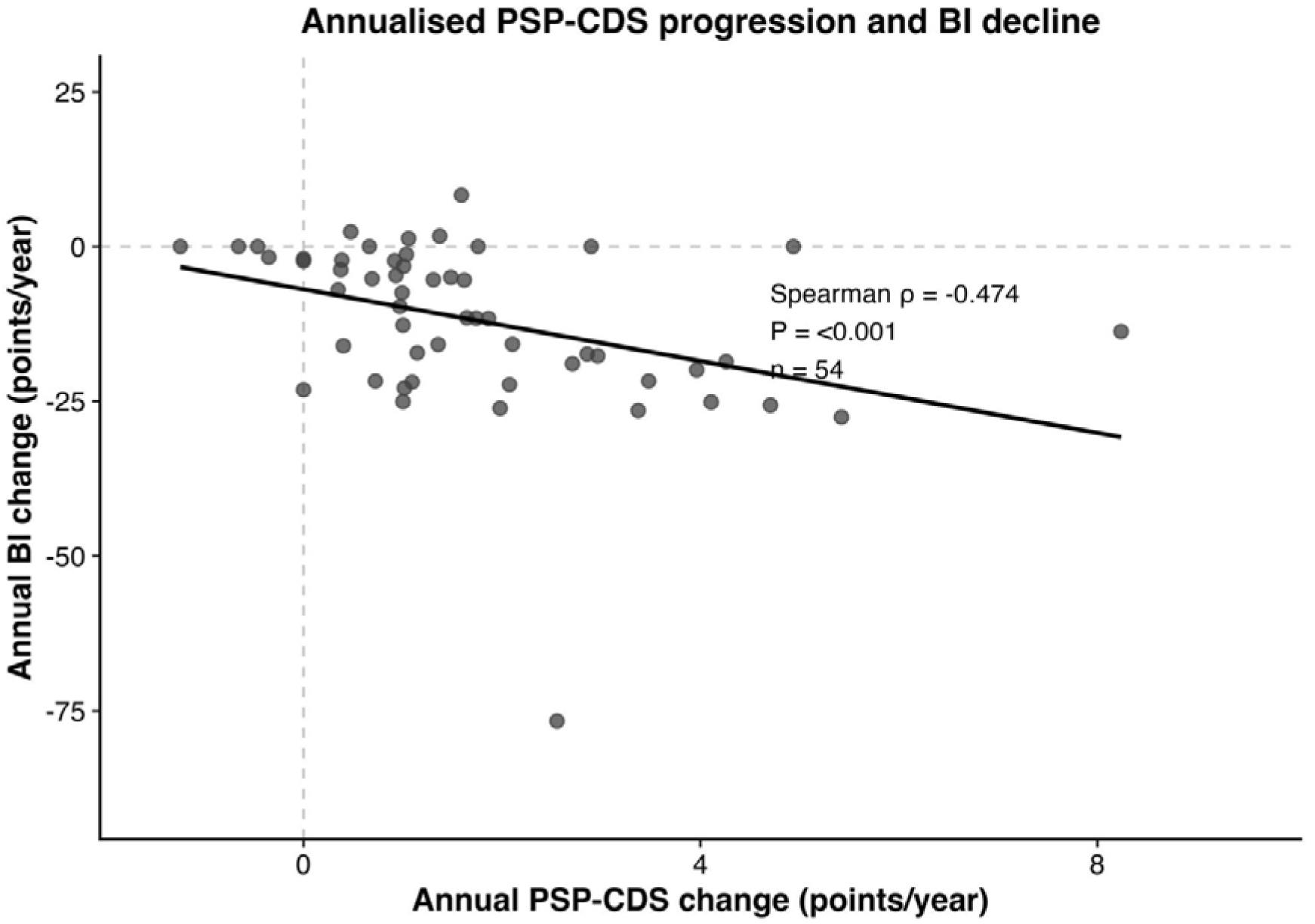
Correlation between annualized changes in PSP-CDS and Barthel Index scores in the overall cohort (Spearman’s rank correlation). Patients with faster worsening of PSP-CDS experienced greater declines in functional independence.

### Power Calculations for Clinical Trials using the PSP-CDS

Table 3 presents the estimated sample sizes required for 9-, 12-, and 18-month clinical trials using the PSP-CDS as the primary outcome measure, assuming a 30% treatment effect, 90% statistical power, and a two-sided significance level of α=0.05. Sample size estimates were based on subtype-specific progression rates and corresponding residual standard deviations derived from the baseline-adjusted linear mixed-effects models. Supplementary Table 4 presents the corresponding sample size estimates under different participant retention scenarios. Sample size estimates suggested that PSP-CDS would be most trial-efficient in PSP-RS and with longer follow-up durations. For 18-month trials, the estimated total sample sizes required were 66, 148, 326, and 646 participants for PSP-RS, the overall cohort, PSP-PGF, and PSP-P, respectively. In comparison, the corresponding total sample sizes required for 9-month trials were 256, 582, 1298, and 2572 participants, respectively. Reduced participant retention substantially increased the required sample sizes across all trial durations and PSP subtypes (Supplementary Table 4).

**Table 3:**
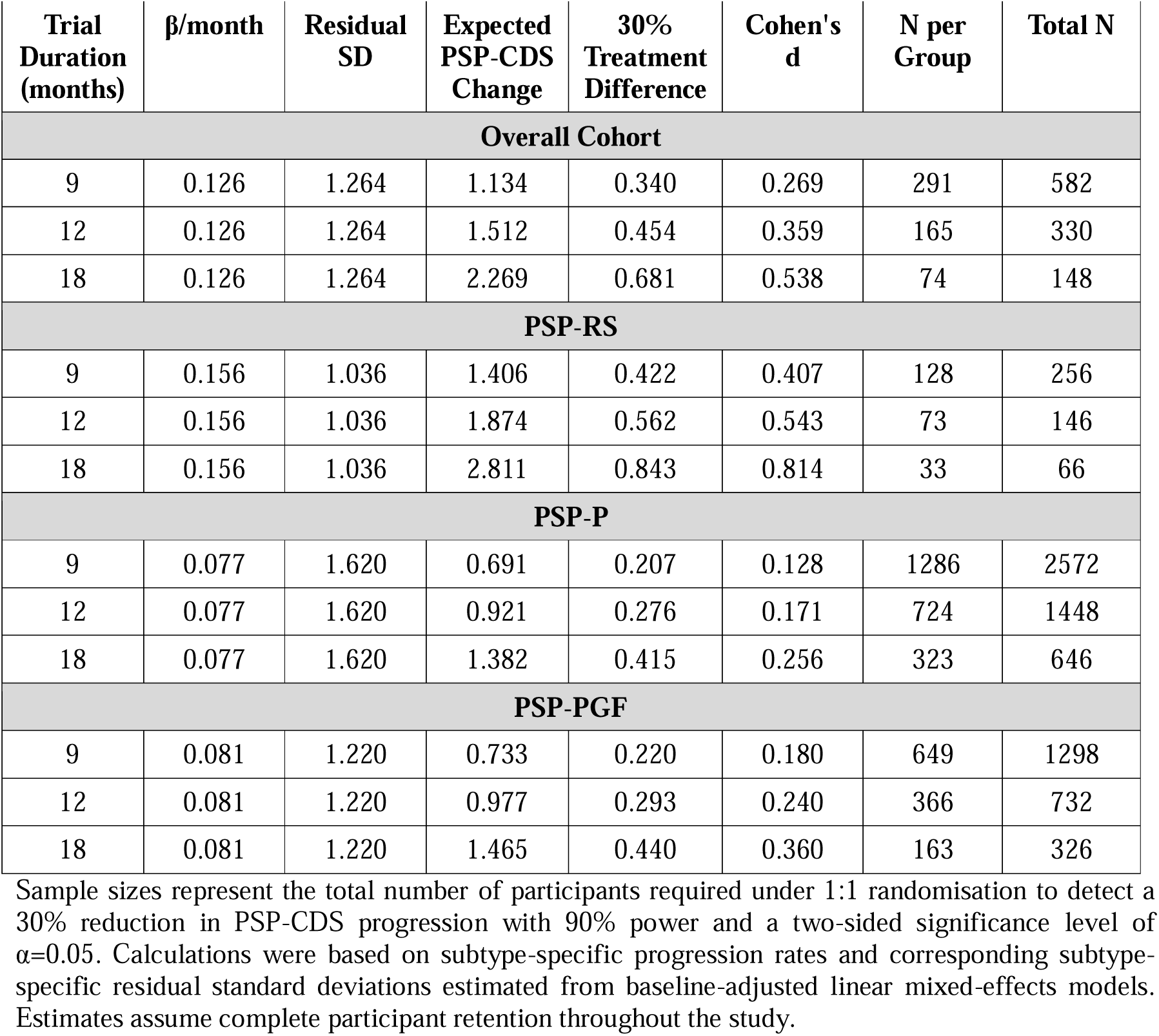
Power calculations for clinical trials using the PSP-CDS.

| **Trial Duration (months)** | **$\beta$/month** | **Residual SD** | **Expected PSP-CDS Change** | **30% Treatment Difference** | **Cohen's d** | **N per Group** | **Total N** |
| --- | --- | --- | --- | --- | --- | --- | --- |
| **Overall Cohort** |  |  |  |  |  |  |  |
| 9 | 0.126 | 1.264 | 1.134 | 0.340 | 0.269 | 291 | 582 |
| 12 | 0.126 | 1.264 | 1.512 | 0.454 | 0.359 | 165 | 330 |
| 18 | 0.126 | 1.264 | 2.269 | 0.681 | 0.538 | 74 | 148 |
| **PSP-RS** |  |  |  |  |  |  |  |
| 9 | 0.156 | 1.036 | 1.406 | 0.422 | 0.407 | 128 | 256 |
| 12 | 0.156 | 1.036 | 1.874 | 0.562 | 0.543 | 73 | 146 |
| 18 | 0.156 | 1.036 | 2.811 | 0.843 | 0.814 | 33 | 66 |
| **PSP-P** |  |  |  |  |  |  |  |
| 9 | 0.077 | 1.620 | 0.691 | 0.207 | 0.128 | 1286 | 2572 |
| 12 | 0.077 | 1.620 | 0.921 | 0.276 | 0.171 | 724 | 1448 |
| 18 | 0.077 | 1.620 | 1.382 | 0.415 | 0.256 | 323 | 646 |
| **PSP-PGF** |  |  |  |  |  |  |  |
| 9 | 0.081 | 1.220 | 0.733 | 0.220 | 0.180 | 649 | 1298 |
| 12 | 0.081 | 1.220 | 0.977 | 0.293 | 0.240 | 366 | 732 |
| 18 | 0.081 | 1.220 | 1.465 | 0.440 | 0.360 | 163 | 326 |

Sample sizes represent the total number of participants required under 1:1 randomisation to detect a 30% reduction in PSP-CDS progression with 90% power and a two-sided significance level of $\alpha=0.05$ . Calculations were based on subtype-specific progression rates and corresponding subtype-specific residual standard deviations estimated from baseline-adjusted linear mixed-effects models. Estimates assume complete participant retention throughout the study.

## DISCUSSION

In this longitudinal study of 104 patients with PSP, who contributed 394 PSP-CDS assessments, we demonstrated that the PSP-CDS is sensitive to longitudinal disease progression in a real-world clinical practice setting. PSP-CDS scores increased significantly over time, with an estimated progression rate of 0.126 points per month, corresponding to approximately 1.13 points over 9 months and 2.27 points over 18 months. Furthermore, and consistent with the recognized clinico-biological heterogeneity of PSP and its subtypes [8–10], subtype-specific analyses demonstrated the fastest progression in PSP-RS, followed by PSP-PGF and PSP-P. PSP-CDS progression was also associated with functional decline, supporting the clinical relevance of the PSP-CDS as a longitudinal outcome measure.

Reliable longitudinal outcome measures are essential for clinical monitoring and research, including studies aimed at understanding the heterogeneous clinical manifestations and progression in PSP. Although the Progressive Supranuclear Palsy Rating Scale (PSPRS) remains the most widely used disease-specific outcome measure, its administration requires considerably more time and expertise than the PSP-CDS. In contrast, the PSP-CDS was specifically developed as a brief and pragmatic tool that can be readily incorporated into routine clinical consultations [13], as was done in this study. Our findings extend previous validation studies by demonstrating that PSP-CDS is not only reflective of disease severity cross-sectionally [5,6] but is also responsive to disease progression over time. Importantly, clinically measurable change was detectable within 9 months, a timeframe highly relevant to observational studies and, potentially, therapeutic trials. To our knowledge, this is the first publication to report on the longitudinal characteristics of the PSP-CDS, and future cross-validation across global cohorts will inform the optimal use of the scale.

Our previous cross-sectional work showed a strong correlation (Spearman’s ρ=-0.81) between the PSP-CDS and the BI [6]. The BI is a generic scale that has been in clinical and research use for ∼60 years, and is widely employed to evaluate patients’ abilities to perform basic physical activities of daily living (ADLs). Its accuracy in measuring physical functionality has been well established in elderly patients and those with disabling diseases such as Parkinson’s disease and stroke. Here, we further showed that the annualized worsening in PSP-CDS scores correlated significantly with annualized declines in BI scores, indicating that patients with faster PSP-CDS progression also experienced greater loss of independence in activities of daily living. These observations indicate that the PSP-CDS assesses functionally relevant clinical features and reinforce its utility as a patient-centered outcome measure [6,13].

One of the main findings of the present study is the heterogeneity in longitudinal progression across PSP phenotypes. PSP-RS demonstrated the fastest progression, followed by PSP-PGF and PSP-P. These observations are consistent with the recognized clinical heterogeneity of PSP and support previous reports that PSP-RS generally follows a more aggressive disease course than variant PSP syndromes, particularly the subcortical forms [8–10]. There have been very limited publications on systematic delineation of PSP subtypes in Asian populations [7,14], and on their influence on PSP progression. Importantly, failure to account for such phenotypic differences may obscure treatment effects in clinical trials and can confound comparisons between cohorts [10]. We and others have highlighted significant differences in Asian populations, in terms of disease epidemiology, clinical presentation, and underlying genetic and environmental factors in PSP, PD, and miscellaneous other movement disorders [5,15–22]

Our exploratory domain-level analyses of the PSP-CDS provide further insight into the patterns of clinical deterioration captured by the scale. Finger dexterity demonstrated the greatest rate of worsening, followed by communication, dysphagia, gait and balance, and eye movements, whereas bradyphrenia and akinesia-rigidity progressed more slowly, with the latter showing minimal progression in PSP-P. We speculate that this may reflect the relatively greater treatment responsiveness of Parkinsonian features compared with the axial motor, ocular motor, and cognitive manifestations of PSP. Notably, several of the most rapidly progressing domains are associated with substantial impairments in communication, swallowing, mobility, and manual dexterity, all of which have an important impact on quality of life, caregiver burden, nutritional status, and clinical outcomes including hospitalization and mortality [23]. These findings suggest that the PSP-CDS captures clinically meaningful aspects of disease progression rather than merely reflecting motor disability. Furthermore, the distinct domain-level progression patterns observed across PSP phenotypes highlight the multidimensional nature of PSP and support the use of the PSP-CDS for monitoring phenotype-specific disease trajectories.

Several study limitations should be considered. There may be selection bias, since our study was conducted at university-based clinics and the longitudinal analyses included only patients with repeated PSP-CDS assessments. However, comparison of baseline characteristics between patients included and excluded from the longitudinal analyses demonstrated similar demographic and clinical characteristics (Supplementary Table 1). Furthermore, since these clinics encompass both public (including free-to-access) and private systems, patients were recruited from diverse socio-demographic backgrounds. Although the overall cohort was substantial for a longitudinal PSP study, the numbers of patients with the less common PSP phenotypes remained relatively small, limiting the precision of subtype-specific estimates. The exploratory residual standard deviation threshold used to estimate detectable progression has not been clinically validated and should therefore be interpreted as an approximate framework for comparing the relative sensitivity of the PSP-CDS across PSP phenotypes rather than as a definitive measure of clinically meaningful change. We did not perform parallel longitudinal analyses using the PSPRS, given the time and human resource constraints in our underserved setting, and the overarching objective of evaluating all patients to maximize research inclusiveness and representativeness [3,17]. The PSPRS, however, remains the most extensively validated disease-specific outcome measure in PSP, and whether the PSP-CDS is sensitive enough to detect small-to-moderate differences in clinical trials of putative disease-modifying agents remains to be seen [24].

On the positive side, this study represents one of the largest longitudinal PSP-CDS cohorts reported from routine clinical practice, comprising almost 400 PSP-CDS assessments. The follow-up duration was relatively long (median almost 3 years, and >5 years in ∼10% of patients), enabling modelling of disease trajectories across multiple repeated measurements, rather than relying solely on single difference scores derived from baseline-to-last follow-up assessments.

In conclusion, the PSP-CDS is a valuable tool for quantifying the severity of PSP and its major subtypes, both cross-sectionally and longitudinally. Besides being useful in routine clinical care, scales like this facilitate accurate documentation of phenotype and clinical severity in Parkinsonian disorders, which serves as a foundation for further studies, including research into the biological factors driving the development and progression of these disabling diseases [25–27]. These efforts can now also be leveraged within collaborative networks harmonizing large datasets, such as the Global Parkinson’s Genetics Program (GP2), which has a strong focus on underrepresented populations and has also recently expanded to include the Parkinson-plus syndromes [17,22,28].

### CRediT AUTHORSHIP CONTRIBUTION STATEMENT

Jun Ping Chua: Writing – review & editing, Writing – original draft, Formal analysis, Data curation. Tzi Shin Toh: Writing – review & editing, Writing – original draft, Supervision, Formal analysis. Ai Huey Tan: Writing – review & editing, Project administration, Investigation. Hirotaka Iwaki: Writing – review & editing, Formal analysis. Shen-Yang Lim: Writing – review & editing, Writing – original draft, Supervision, Project administration, Investigation, Data curation, Conceptualization. All other authors: Writing – review & editing.

## Supporting information

GP2 banner author list

## ACKNOWLEDGEMENTS

The authors gratefully acknowledge the generous participation of patients and their caregivers at University of Malaya. This work was supported by the University of Malaya Parkinson’s Disease and Movement Disorders Research Program (PV035-2017) awarded to SYL and AHT.

## DATA AVAILABILITY

The clinical data used in the preparation of this article will be included in a future data release from the Global Parkinson’s Genetics Program (GP2; https://gp2.org), with corresponding genetics data. Specifically, we will share Tier 2 data in GP2 release 13 (see current releases at https://zenodo.org/records/20932193). GP2 data can be requested through AMP PD (https://amp-pd.org).

## CODE AVAILABILITY

All code generated for this article, and the identifiers for all software programs and packages used, are available on Github (https://github.com/GP2code/mys_psp-cds) and were given a persistent identified via Zenodo (10.5281/zenodo.21481788).

