## Supplementary Materials for "The Progressive Supranuclear Palsy Clinical Deficits Scale (PSP-CDS) tracks longitudinal progression in PSP"

**Supplementary Table 1: Comparison of baseline clinico-demographic characteristics between patients included and excluded from the longitudinal analyses.**

|  | **Study Cohort** | **Excluded Cohort** | **P-value** |
| --- | --- | --- | --- |
| **N** | 104 (100.0) | 76 (73.1) | - |
| **Sex**  Male  Female | 61 (58.7)  43 (41.3) | 47 (61.8)  29 (38.2) | 0.782 |
| **Clinical PSP subtype**  PSP-RS  PSP-P  PSP-PGF  Other PSP variants | 59 (56.7)  28 (26.9)  14 (13.5)  3 (2.9) | 37 (48.7)  20 (26.3)  11 (14.5)  8 (10.5) | 0.193 |
| **Ethnicity**  Chinese  Indian  Malay  Others | 73 (70.2)  20 (19.2)  9 (8.7)  2 (1.9) | 45 (59.2)  18 (23.7)  10 (13.2)  3 (3.9) | 0.443 |
| **Age at symptom onset** (years)  **Age at baseline** (years)  **Disease duration at baseline** (years)  **PSP-CDS at baseline** | 67.0 [61.0-72.0]  72.1 [65.9-76.5]  4.5 [2.6-6.4]  9.0 [7.0-12.0] | 68.0 [62.0-72.0]  72.7 [65.9-76.8]  4.5 [2.5-6.4]  10.0 [7.0-14.2] | 0.444  0.503  0.889  0.044* |

Categorical variables are presented as n (%), and continuous variables are presented as median [Q1–Q3].

P-values were calculated using Pearson’s chi-square test or Fisher’s exact test for categorical variables, as appropriate, and the Kruskal–Wallis rank-sum test for continuous variables.

*Denotes statistically significant differences (P<0.05).

PSP=Progressive supranuclear palsy; PSP-CDS=PSP Clinical Deficits Scale; PSP-PGF=PSP with progressive gait freezing; PSP-P=PSP with predominant parkinsonism; PSP-RS=PSP Richardson’s syndrome.

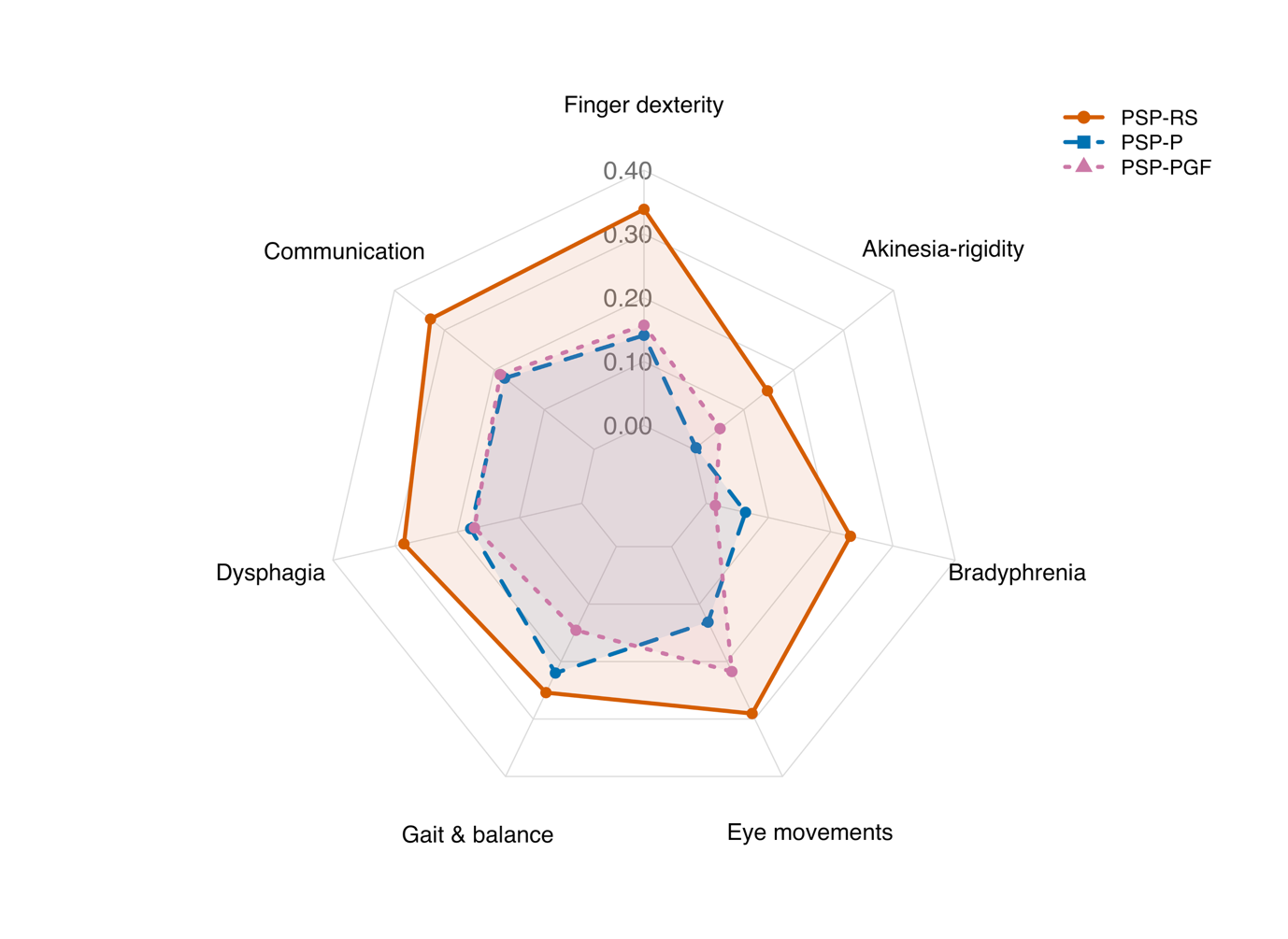

**Supplementary Figure 1: Estimated annual progression of individual PSP-CDS domains across major PSP subtypes.** Radar chart illustrating estimated annual changes (β/year) in individual PSP-CDS domains derived from subtype-specific baseline-adjusted linear mixed-effects models incorporating the corresponding baseline domain score, age at baseline, and sex. Greater radial distance indicates greater estimated annual progression. Domains are ordered according to the magnitude of annual progression observed in the overall cohort to facilitate comparison across PSP subtypes. PSP-RS demonstrated the greatest progression across most domains, particularly finger dexterity, communication, dysphagia, gait and balance, and eye movements, whereas PSP-P and PSP-PGF exhibited lower overall progression with distinct domain-specific patterns of progression.

**Supplementary Table 2 : Estimated annual progression of individual PSP-CDS domains in the overall cohort and across PSP subtypes.**

| **Domain** | **Overall Cohort** | **PSP-RS** | **PSP-P** | **PSP-PGF** |
| --- | --- | --- | --- | --- |
| Finger dexterity | 0.264 (0.201–0.327); P<0.001 | 0.339 (0.246–0.432); P<0.001 | 0.141 (0.063–0.220); P=0.002 | 0.157 (0.045–0.269); P=0.018 |
| Communication | 0.262 (0.203–0.321); P<0.001 | 0.328 (0.249–0.407); P<0.001 | 0.179 (0.080–0.278); P=0.002 | 0.188 (0.039–0.338); P=0.028 |
| Dysphagia | 0.244 (0.183–0.304); P<0.001 | 0.286 (0.193–0.379); P<0.001 | 0.179 (0.082–0.276); P=0.001 | 0.172 (0.049–0.296); P=0.018 |
| Gait & balance | 0.226 (0.178–0.275); P<0.001 | 0.254 (0.187–0.321); P<0.001 | 0.220 (0.121–0.319); P<0.001 | 0.146 (0.037–0.254); P=0.023 |
| Eye movements | 0.221 (0.161–0.281); P<0.001 | 0.290 (0.221–0.360); P<0.001 | 0.131 (0.020–0.242); P=0.027 | 0.217 (0.074–0.360); P=0.010 |
| Bradyphrenia | 0.145 (0.071–0.220); P<0.001 | 0.232 (0.112–0.352); P<0.001 | 0.063 (-0.053–0.179); P=0.295 | 0.015 (-0.097–0.126); P=0.802 |
| Akinesia-rigidity | 0.093 (0.033–0.153); P=0.003 | 0.147 (0.058–0.237); P=0.002 | 0.004 (-0.097–0.105); P=0.935 | 0.052 (-0.080–0.185); P=0.452 |

The overall analysis included 394 observations from 104 participants. Subtype-specific analyses included 211 observations from 59 participants with PSP-RS, 107 observations from 28 participants with PSP-P, and 63 observations from 14 participants with PSP-PGF. Values represent estimated annual changes (β/year) with corresponding 95% confidence intervals and exact P-values derived from baseline-adjusted linear mixed-effects models including follow-up time, the corresponding baseline domain score, age at baseline, and sex as fixed effects, with participant-specific random slopes for follow-up time (0 + Time_months | ID). Domains are ranked according to the magnitude of annual progression observed in the overall cohort. Exact P-values correspond to the fixed effect of follow-up time in each model.

**Supplementary Table 3: Time intervals between matched PSP-CDS and BI assessments at baseline and follow-up.**

|  | Baseline, n (%) | Follow-up, n (%) |
| --- | --- | --- |
| Same day | 48 (88.9) | 41 (75.9) |
| 1–30 days | 2 (3.7) | 0 (0.0) |
| 31–90 days | 1 (1.9) | 2 (3.7) |
| 91–180 days | 3 (5.6) | 4 (7.4) |
| >180 days | 0 (0.0) | 7 (13.0) |

**Supplementary Table 4: Estimated sample sizes required for clinical trials under different participant retention scenarios.**

| **Trial Duration (months)** | **100% Retention** | **90% Retention** | **80% Retention** | **70% Retention** | **60% Retention** |
| --- | --- | --- | --- | --- | --- |
| **Overall Cohort** | | | | | |
| 9 | 582 | 648 | 728 | 832 | 970 |
| 12 | 330 | 368 | 414 | 472 | 550 |
| 18 | 148 | 166 | 186 | 212 | 248 |
| **PSP-RS** | | | | | |
| 9 | 256 | 286 | 320 | 366 | 428 |
| 12 | 146 | 164 | 184 | 210 | 244 |
| 18 | 66 | 74 | 84 | 96 | 110 |
| **PSP-P** | | | | | |
| 9 | 2572 | 2858 | 3216 | 3676 | 4288 |
| 12 | 1448 | 1610 | 1810 | 2070 | 2414 |
| 18 | 646 | 718 | 808 | 924 | 1078 |
| **PSP-PGF** | | | | | |
| 9 | 1298 | 1444 | 1624 | 1856 | 2164 |
| 12 | 732 | 814 | 916 | 1046 | 1220 |
| 18 | 326 | 364 | 408 | 466 | 544 |

Sample sizes represent the total number of participants required under 1:1 randomization to detect a 30% reduction in PSP-CDS progression with 90% power and a two-sided significance level of α=0.05. Calculations were based on subtype-specific progression rates and corresponding subtype-specific residual standard deviations estimated from baseline-adjusted linear mixed-effects models.
